# Fluid and Neuroimaging Biomarkers in Microgliopathy Colony-Stimulating Factor-1 Receptor-Related Disorders

**DOI:** 10.1101/2025.09.12.25335644

**Authors:** Tomasz Chmiela, Madison Reeves, Karen Jansen-West, Judith Dunmore, Yuping Song, Audrey Strongosky, Sunil Gandhi, Gilana Pikover, Matt Blurton-Jones, Robert C. Spitale, Erik H. Middlebrooks, Leonard Petrucelli, Mercedes Prudencio, Zbigniew K. Wszolek

**Affiliations:** Department of Neurology, Mayo Clinic, Jacksonville; 4500 San Pablo Rd S, 32224 FL, USA; Department of Neurology, Faculty of Medical Sciences, Medical University of Silesia; Medyków 14, 42-752 Katowice, Poland; Department of Neuroscience, Mayo Clinic, Jacksonville; 4500 San Pablo Rd S, 32224 FL, US; Savanna Biotherapeutics, Inc.; 1 University Drive, Aliso Viejo, CA 92656; Department of Radiology, Mayo Clinic; 4500 San Pablo Road, Jacksonville, FL 32224, USA; Neurobiology of Disease Graduate Program, Mayo Graduate School, Mayo Clinic College of Medicine, Jacksonville, FL, 32224, USA

## Abstract

Colony-stimulating factor 1 receptor-related disorder (CSF1R-RD) is a neurodegenerative condition characterized by rapid progression, leading to profound functional decline and ultimately resulting in a persistent vegetative state. Although an effective treatment option exists, there remains a lack of identified biomarkers capable of monitoring disease progression and detecting the earliest symptom onset in *CSF1R* pathogenic variant carriers, limiting the ability of clinicians to make informed decisions regarding patient care. This study aims to identify both fluid and neuroimaging biomarkers for CSF1R-RD that can inform the optimal timing of treatment administration to maximize therapeutic benefit, while also providing sensitive quantitative measurements to monitor disease progression. Our study compared neuroimaging and fluid (plasma and cerebrospinal fluid (CSF)) biomarkers across three distinct populations: asymptomatic *CSF1R* pathogenic variant carriers (N=14), symptomatic *CSF1R* pathogenic variant carriers (N=17), and healthy controls (N=30). We evaluated biomarker correlations with both an established (Montreal Cognitive Assessment (MoCA)) and a novel (CSF1R Clinical Severity Score (CCSS)) clinical diagnostic scale to investigate potential clinical utility. Additionally, we tested the relationship between select biomarkers and cortical thickness using 3D T1-weighted MPRAGE scans, providing a highly valuable physiological component to our analyses. Our results demonstrate that while plasma glial fibrillary acidic protein (GFAP) displays a high sensitivity for distinguishing early-stage CSF1R-RD patients from healthy controls, plasma neurofilament light chain (NfL) is more effective for tracking disease progression following the onset of symptoms. Overall, our study provides evidence for plasma NfL and GFAP as valuable biomarkers of earliest symptom onset and disease progression for CSF1R-RD

**One Sentence Summary:** This study identifies plasma biomarkers NfL and GFAP as promising tools to detect CSF1R-RD onset and progression, with potential to improve patient outcomes.

## INTRODUCTION

Colony stimulating factor 1 receptor (CSF1R)-related disorder (CSF1R-RD) is a rare, hereditary, and rapidly progressive neurodegenerative disease caused by pathogenic variants in *CSF1R* gene ^1^. The *CSF1R* gene encodes a tyrosine kinase transmembrane receptor involved in the survival, proliferation, and differentiation of microglia ^2, 3^. The causative role of *CSF1R* variants in adult-onset leukoencephalopathy with axonal spheroids and pigmented glia was identified in 2011 ^4^. Most pathogenic variants, located in the tyrosine kinase domain (TKD, exons 12–22) ^5, 6^, cause the downregulation of autophosphorylation and the inactivation of tyrosine kinases, thereby influencing the downstream signaling pathway ^4^. The growing availability of genetic testing has led to increased recognition and diagnosis of CSF1R-RD. CSF1R-RD has an estimated global prevalence of 0.5-1.5 per 100,000, with cases reported in the Americas, Asia, Australia, and Europe ^7, 8^. According to a recent study by Wade et al., which analyzed the prevalence of pathogenic and likely pathogenic variants of *CSF1R* gene in the British BioBank, the prevalence of these variants may even be as high as 28.09/100,000 ^9^.

Pathological variants in the *CSF1R* gene cause primary microgliopathy that affects axon integrity leading to progressive and eventually fatal leukoencephalopathy. Pathologically, CSF1R-RD is characterized by demyelination of cerebral white matter, frontoparietal lobe atrophy, thinning of the corpus callosum, swelling of axons, and pigmented glial cells ^4^.

Diagnostic criteria for CSF1R-RD, established in 2017, aids in identifying patients with potential CSF1R-RD; however, a definitive diagnosis requires the identification of a pathogenic variant in *CSF1R* gene ^10^. This makes diagnosing CSF1R-RD challenging, with only 25% of affected patients initially receiving the correct diagnosis, with the most common misdiagnoses being frontotemporal dementia, multiple sclerosis, or non-specific neurodegeneration/dementia ^11^.

Clinically, CSF1R-RD manifests as personality and behavioral changes, dementia, and motor symptoms. The age of onset ranges from early onset (<18 years of age) to late onset (≥18 years of age), with a mean age of symptom onset of 40±10 years in women and 47±11 years in men ^1^. In most of the affected individuals, rapid progression of disability is observed, leading very quickly to total incapacitation ^1, 7^. Besides the progression of motor symptoms, patients lose their ability to speak, perform voluntary movements, and eventually appear unaware of their surroundings. In the late stage, symptoms progress to a vegetative state with the presence of primitive reflexes ^7, 10, 11^. Despite significant progress in understanding the clinical characteristics of CSF1R-RD, no unified clinical scale currently exists that comprehensively captures the full spectrum of symptoms associated with the condition. Rating patients in the early stages can be especially challenging, as they may present with diverse clinical phenotypes, with dominance of motor or cognitive symptoms.

With a growing understanding of the pathophysiology of CSF1R-RD ^1, 12^, therapeutic options are emerging, including the promising results of off-label hematopoietic stem cell transplantation (HSCT) ^13, 14^. Another promising option for asymptomatic *CSF1R* pathogenic variant carriers might be corticosteroid prophylaxis. This consideration is supported by observational data indicating that individuals undergoing long-term corticosteroid treatment for unrelated conditions (e.g., asthma, rheumatoid arthritis) tend to exhibit a delayed onset of CSF1R-RD symptoms ^12, 15^. This protective effect has been corroborated in a CSF1R-RD animal model ^16^.

Neuroimaging plays a crucial role in the diagnosis of CSF1R-RD and often prompts genetic testing in affected individuals. Typical radiologic findings in magnetic resonance imaging (MRI) include progressive bilateral white matter lesions (hyperintense on T2-weighted and FLAIR sequences, hypointense on T1-weighted images) that are patchy and focal in the early stages and become more confluent as the disease progresses cerebral atrophy ^17, 18^; and thinning of the corpus callosum. White matter calcifications have been observed in half of CSF1R-RD ^5^ In 2012, Sundal et al. ^19^ developed a scale to assess the severity of changes in MRI dedicated to CSF1R-RD. However, it is still unclear whether neuroimaging can be helpful in detecting conversion to the symptomatic stage and whether it can be an efficient method for monitoring disease progression.

Therefore, there is a growing interest in identifying precise and objective biomarkers (including neuroimaging and laboratory tests) that would allow for increased diagnostic sensitivity to help monitor patient status and to serve as a reference for evaluating treatment trials. The optimal way to assess these patients has not been determined despite advances in our understanding of CSF1R-RD. The identification of biomarkers in bodily fluids is a particularly desirable solution due to its reliability, repeatability, and minimal invasiveness.

To address the current lack of robust biomarker data and the critical need for reliable diagnostic markers of disease progression, we leveraged a large cohort of *CSF1R* pathogenic variant carriers to a conduct a comprehensive analysis of fluid biomarkers (NfL, macrophage colony stimulating factor (M-CSF), glial fibrillary acidic protein (GFAP), interleukin-34 (IL-34) and osteopontin, in both serum and CSF), neuroimaging biomarkers (MRI), a novel clinical diagnostic scale tool [CSF1R Clinical Severity Score (CCSS)] and the Montreal Cognitive Assessment clinical (MoCA) test. We assessed the potential of these biomarkers to reflect clinically detectable disease progression. We also developed a novel clinical diagnostic scale designed to capture the heterogenous nature of CSF1R-RD and to serve as a standardized and highly sensitive metric of clinical symptoms associated with CSF1R-RD.

## RESULTS

### Study cohort

We leveraged a robust cohort of 17 symptomatic and 14 asymptomatic *CSF1R* pathogenic variant carriers and 30 healthy controls (N = 61) to identify candidate biomarkers capable of guiding the optimal timing of therapeutic intervention for presymptomatic carriers transitioning to symptomatic states, and enable the sensitive monitoring of disease progression after symptom onset. The 31 *CSF1R* pathogenic carriers (11 males, 35.5%; 20 females, 64.5%) had a mean age of 47.9±15.9 years (range 20-77 years). Individuals were identified as asymptomatic (mean age: 46.1±17.5 years, range 22-73 years) based on the presence of a genetically confirmed *CSF1R* pathogenic variant and no detectable clinical symptoms associated with CSF1R-RD. Individuals were classified as symptomatic (mean age: 49.4±14.8 years) given the presence of a pathogenic variant along with a clinically-detectable symptom onset (mean age of symptom onset: 43.5±16.0 years), ranging from the presence of cognitive symptoms, mood impairment, behavioral changes, motor symptoms like parkinsonism, cerebellar and pyramidal signs, and sensory dysfunction. In the studied cohort, 11 out of 17 individuals (64.7%) exhibited cognitive symptoms, ranging from mild cognitive impairment to dementia. Behavioral changes were observed in 7 individuals (41.2%). Motor symptoms were present in 14 individuals (82.4%), including parkinsonism (70.5%), increased muscle tone (76.5%)—typically a combination of spasticity and rigidity—along with pyramidal (58.8%) and cerebellar signs (47.1%) and gait abnormalities (76.5%).

A list of all *CSF1R* pathogenic variants detected in this cohort is provided in **Supplementary Figure 1**. The control group consisted of 30 healthy negative for *CSF1R* pathogenic variants (18 males, 60%; 12 females, 40%) with a mean age of 51.4±13.0 years (range 32-75 years). Twenty-three participants with *CSF1R* pathogenic variant carriers (13 asymptomatic, and 10 symptomatic) had 3-Tesla MRI of the brain available, obtained within one week of biomarker collection.

### CSF1R-RD patients show significant white matter atrophy

Clinical diagnostic severity was calculated from neuroimaging MRI scans using the Sundal scale ^19^ (**Table 2**). Symptomatic patients were characterized by significantly higher total Sundal scale scores [11 (6.5, 13.5) points] compared to asymptomatic [3 (0, 4) points] individuals (**Figure 1A**, **Table 2**). Analysis of individual Sundal subscoring components showed that the atrophy component was significantly elevated in symptomatic patients [4 (0.5, 4) points] compared to asymptomatic individuals [0 (0, 0) points] (**Supplementary Figure 2A**, **Table 2**). Notably, the white matter subscoring component was even more significantly elevated symptomatic patients [7 (6.5, 9.5) points] when compared with asymptomatic individuals [3 (0, 4 points] (**Supplementary Figure 2B**, **Table 2**). Symptomatic patients also exhibited substantially higher white matter lesion volume [20995 (4418, 38488) mm^3^] compared to asymptomatic patients [179 (12, 573) mm^3^] (**Figure 1B**, **Table 2**). Multiple neuroimaging markers showed trends toward reductions in symptomatic patients compared to asymptomatic patients, including normalized brain volume [0.73 (0.69, 0.79) vs. 0.79 (0.78, 0.81) mm³; p=0.034] (**Table 2, Supplementary Figure 2C**), normalized white matter volume [0.27 (0.26, 0.32) vs. 0.32 (0.30, 0.32) mm³; p=0.077] (**Table 2, Supplementary Figure 2D**), corpus callosum volume [2568.6 (1693.2, 3471.2) vs. 3528.5 (3372.3, 3810.8) mm³; p=0.057] (**Table 2, Supplementary Figure 2E**), cerebral white matter volume [421.6 (388.5, 453.2) vs. 481.3 (433.9, 503.4) cm³; p=0.166] (**Table 2, Supplementary Figure 2F**), subcortical gray matter volume [52.8 (49.8, 56.2) vs. 60.0 (57.6, 61.3) cm³; p=0.067] (**Table 2, Supplementary Figure 2G**), and ventricular volume [56.1 (39.9, 68.8) vs. 20.8 (18.2, 27.3) cm³; p=0.015] (**Table 2, Supplementary Figure 2H**). However, none of these differences reached the Bonferroni-corrected significance threshold (p<0.0045). Cortex volume did not show any substantial difference between symptomatic and asymptomatic patients [(458.0 (446.6, 521.2) vs. 500.1 (462.9, 532.7) cm³; p=0.483] (**Table 2, Supplementary Figure 2I**).

**Figure 1.**
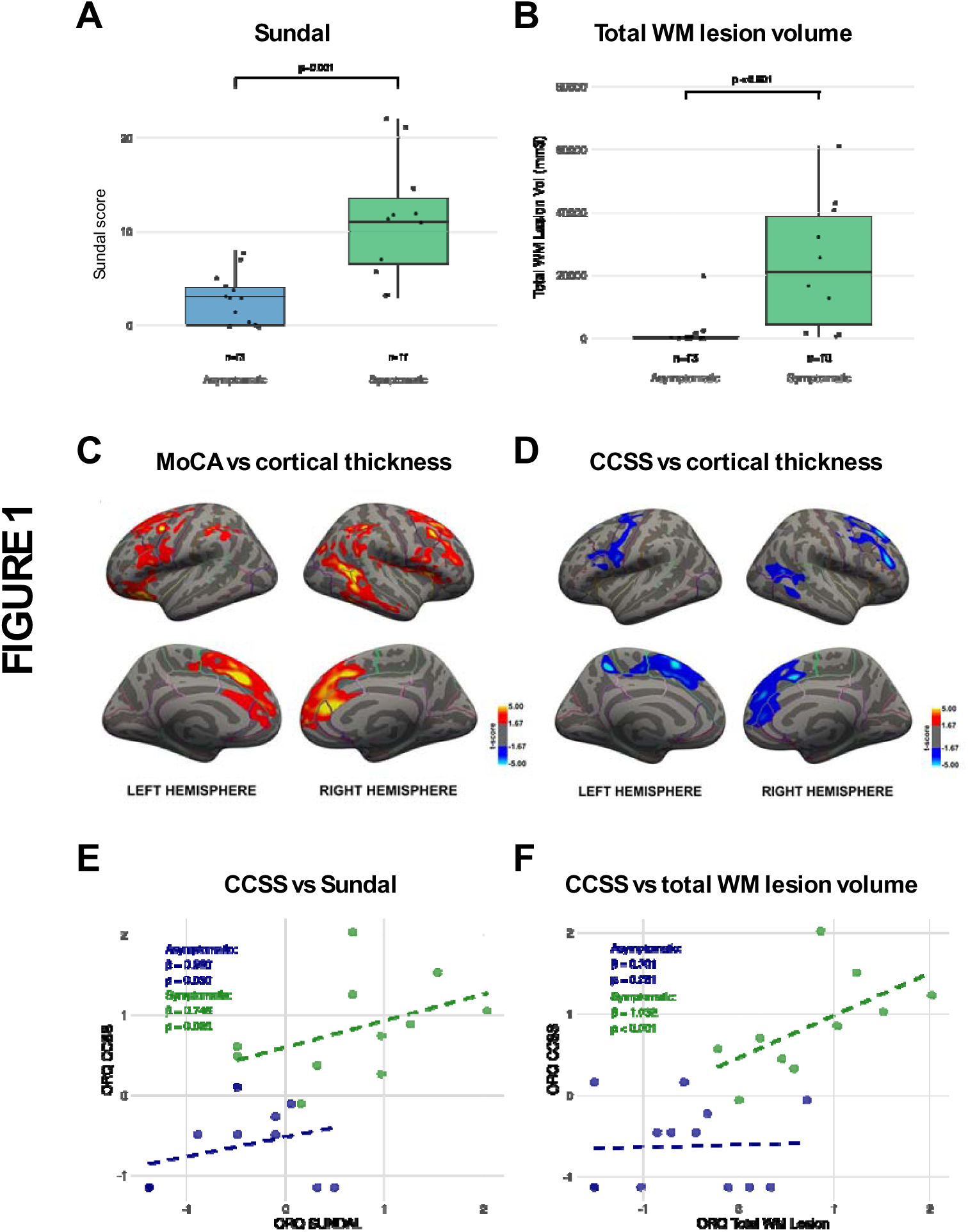
Symptomatic patients reflect higher clinical diagnostic severity and physiological changes. Sundal scores (**A**) and total white matter lesion volume (**B**) are significantly elevated in symptomatic patients compared to asymptomatic patients. Correlations of MoCA (**C**) and CCSS (**D**) with cortical thickness measured with 3D T1-weighted MPRAGE scans across multiple brain regions indicate that both clinical assessments are reflective of the physiological changes associated with CSF1R-RD disease progression. (**E-F**) Scatterplots of ranked relationships (ORQ-transformed) between CSF1R-RD Clinical Severity Score (CCSS) and Sundal scores (E) or total white matter lesion volume (**F**).

### CSF1R-RD patients show changes in MoCA and CCSS

The Montreal Cognitive Assessment (MoCA) is a well-established clinical diagnostic tool used to track the severity of cognitive decline in patients. Patients begin with a score of 30 and lose points as they lose cognitive function. Cognition of symptomatic CSF1R-RD patients was affected in our study cohort with a median score of 15, while asymptomatic individuals were virtually unaffected with a median score of 28 (**Table 1**). To affirm the informative ability of MoCA in CSF1R-RD patients, we obtained cortical thickness measurements using 3D T1-weighted MPRAGE imaging. Higher MoCA scores associated with higher cortical thickness across multiple brain regions, including the bilateral prefrontal regions, cingulum, and inferior parietal lobe, as well as the right lateral temporal lobe (**Figure 1C**). Moreover, higher MoCA scores correlated with lower total white matter lesion volume (r=-0.60, p=0.003), lower Sundal scores (r=-0.51, p=0.012), and higher corpus callosum volume (r=0.53, p=0.011) (**Table 3**). In contrast, normalized brain volumes (r=0.44, p=0.040), subcortical gray matter volume (r=0.34, p=0.118), normalized white matter volume (r=0.37, p=0.087), cortex volume (r=0.19, p=0.393), cerebral white matter volume (r=0.23, p=0.306), ventricle volume (r=-0.46, p=0.032) did not correlate with MoCA scoring (**Table 3**).

**Table 1.** Descriptive statistics of the study cohort.

|  | CSF1R-RD |  |  | Controls<br>N = 30 |
| --- | --- | --- | --- | --- |
|  | All<br>N = 31 | Asymptomatic<br>N = 14 | Symptomatic<br>N = 17 |  |
| Age at biomarker collection [mean±SD (range)] | 47.9±15.9<br>(20.6-77.1) | 46.1±17.5<br>(20.6-77.1) | 49.4±14.8<br>(22.0-72.5) | 51.4±13.0<br>(31.7-75.0) |
| Age at symptom onset<br>[mean±SD (range)] | N/A | N/A | 43.5±16.0<br>(5, 7.1) | N/A |
| Sex [n (%)] |  |  |  |  |
| Male | 11 (35.5) | 5 (35.6) | 6 (35.3) | 18 (60) |
| Female | 20 (64.5) | 9 (64.3) | 11 (64.7) | 12 (40) |
| CCSS Total [median (IQR)] | 5 (0, 29) | 1 (0, 2.8) | 29 (9.5, 47) | N/A |
| CCSS Cognition [median (IQR)] | 0 (0, 6) | 0 (0, 0) | 6 (0, 21.5) | N/A |
| CCSS Affect and Mood [median (IQR)] | 0 (0, 1) | 0 (0, 0) | 1 (0, 5.5) | N/A |
| CCSS Cranial Nerves [median (IQR)] | 0 (0, 0) | 0 (0, 0) | 0 (0, 0) | N/A |
| CCSS Motor [median (IQR)] | 4 (0, 14) | 0 (0, 2) | 14 (6.5, 25) | N/A |
| CCSS Sensory [median (IQR)] | 0 (0, 0) | 0 (0, 0) | 0 (0, 2) | N/A |
| CCSS Other [median (IQR)] | 0 (0, 2) | 0 (0, 0) | 2 (0, 3.5) | N/A |
| MoCA [median, (IQR)] | 27 (15, 29) | 28 (28, 30) | 15 (11, 26.5) | N/A |

CCSS: CSF1R-RD Clinical Severity Score; MoCA: Montreal Cognitive Assessment; IQR: interquartile range; SD: standard deviation; n: number of cases; N/A – non applicable.

**Table 2.** Differences in white matter lesions and atrophy, as well as biofluid biomarkers, between healthy controls, asymptomatic and symptomatic *CSF1R* mutation carriers.

|  | Asymptomatic<br>[Median (IQR)] | Symptomatic<br>[Median (IQR)] | Healthy controls<br>[Median (IQR)] | p-value |
| --- | --- | --- | --- | --- |
| **MRI** |  |  |  |  |
| Sundal scale |  |  |  |  |
| Total | 3 (0-4) | 11 (6.5-13.5) | N/A | **0.001** |
| White matter (WM) | 3 (0-4) | 7 (6.5-9.5) | N/A | **<0.001** |
| Atrophy | 0 (0-0) | 4 (0.5-4) | N/A | 0.007 |
| Total WM lesion volume (mm<sup>3</sup>) | 179 (12, 573) | 20995 (4418,38488) | N/A | **<0.001** |
| Normalized brain volume | 0.79 (0.78, 0.81) | 0.73 (0.69, 0.79) | N/A | 0.034 |
| Normalized WM volume | 0.32 (0.30, 0.32) | 0.27 (0.26, 0.32) | N/A | 0.077 |
| Corpus callosum volume (mm<sup>3</sup>) | 3528.5 (3372.3, 3810.8) | 2568.6 (1693.2, 3471.2) | N/A | 0.057 |
| Cortex volume (cm<sup>3</sup>) | 500.1 (462.9, 532.7) | 458.0 (446.6, 521.2) | N/A | 0.483 |
| Cerebral WM volume (mm<sup>3</sup>) | 481.3 (433.9, 503.4) | 421.6 (388.5, 453.2) | N/A | 0.166 |
| Subcortical gray matter volume (cm<sup>3</sup>) | 60.0 (57.6, 61.3) | 52.8 (49.8, 56.2) | N/A | 0.067 |
| Ventricles volume (cm<sup>3</sup>) | 20.8 (18.2, 27.3) | 56.1 (39.9, 68.8) | N/A | 0.015 |
| **Plasma biomarkers** |  |  |  |  |
| M-CSF (pg/mL) | 29.8 (26.4, 36.3) | 31.4 (26.4, 35.2) | 23.8 (21.0, 27.4) | **0.002** |
| IL-34 (pg/mL) | 2.7 (1.7, 3.1) | 2.5 (2.0, 3.2) | 1.5 (1.1, 1.9) | **<0.001** |
| NfL (pg/mL) | 6.2 (4.5, 13.1) | 38.5 (14.8, 102.9) | 6.6 (4.4, 9.6) | **<0.001** |
| Osteopontin (pg/mL) | 3871.3 (2861.1, 4848.7) | 3327.0 (2481.2, 4351.5) | 4087.8 (3100.2, 4712.7) | 0.540 |
| GFAP (pg/mL) | 181.1 (128.9, 246.5) | 280.0 (177.8, 689.4) | 89.3 (70.5, 154.2) | **<0.001** |
| **CSF biomarkers** |  |  |  |  |
| M-CSF, (pg/mL) | 7.9 (6.8, 10.4) | 13.3 (10.8, 15.8) | 5.5 (4.2, 6.7) | **<0.001** |
| IL 34, (pg/mL) | 142.5 (123.6, 151.6) | 132.3 (95.2, 224.4) | 58.2 (46.1, 71.7) | **<0.001** |
| NfL, (pg/mL) | 505.0 (287.6, 612.9) | 2972.4 (2205.2, 4646.9) | 489.2 (381.6, 612.8) | 0.019 |
| Osteopontin (pg/mL) | 12723.8 (9835.2, 17367.7) | 16578.9 (7117.5, 24452.2) | 16949.1 (8400.4, 21772.9) | 0.797 |
| GFAP (pg/mL) | 12102.5 (9129.2, 14654.7) | 20920.3 (10805.1, 24752.7) | 8405.1 (5628.6, 11631.5) | 0.007 |

Mann-Whitney U tests were employed for MRI level comparisons due to violation of normality assumptions. For Mann-Whitney U tests, p-values < 0.0045 were considered significant after applying a Bonferroni correction for multiple testing. Kruskal-Wallis tests were employed for plasma and CSF level comparisons due to violation of normality assumptions. For Kruskal-Wallis tests, p-values < 0.01 were considered significant after applying a Bonferroni correction for multiple testing. IQR: interquartile range; WM: white matter; CSF: cerebrospinal fluid; M-CSF: macrophage colony stimulating factor; IL-34: interleukin-34; NfL: neurofilament light chain; GFAP: glial fibrillary acidic protein; N/A: not applicable.

**Table 3.**
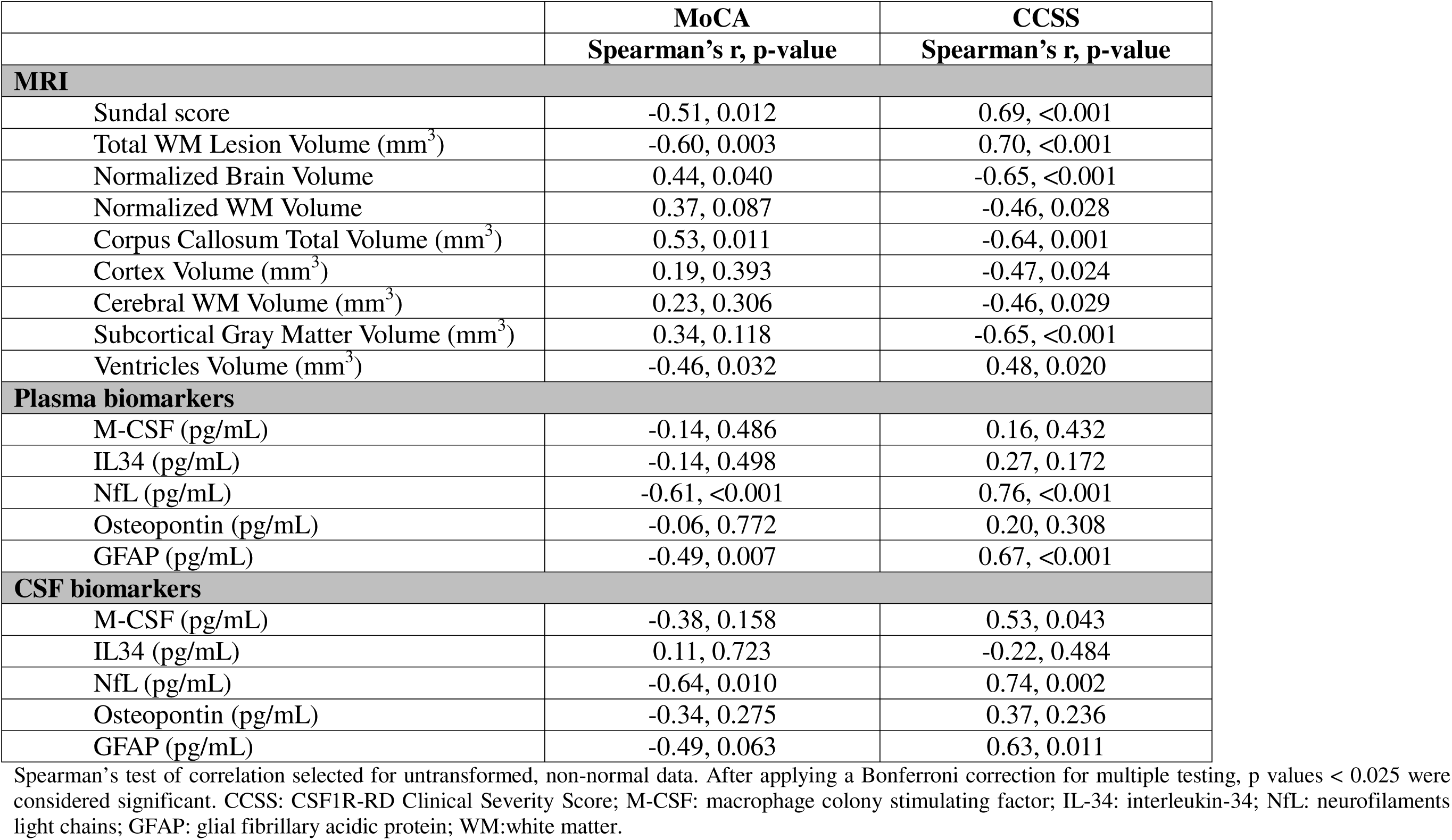
Correlation between brain lesion volume, brain atrophy, fluid biomarker levels and clinical severity diagnostics.

Next, to better evaluate symptom severity in CSF1R-RD patients, we developed the CSF1R Clinical Severity Score (CCSS), designed to maximize sensitivity to symptom progression. The CCSS is a semi-quantitative scale that assesses the patient in five domains: cognition, mood and affect, cranial nerves, motor function and sensory function, and is sensitive enough to detect even minor cognitive and motor deviations. Scores range from 0 (unaffected) to 244 (severe) (**Supplementary Tool 1**). Symptomatic *CSF1R* carriers showed median CCSS total values of 29, scoring most strongly with worsened cognition (4) and motor function (14) (**Table 1**). To determine the utility of the CCSS, we also compared it to the neuroanatomical changes observed using 3D T1-weighted MPRAGE scans. CCSS showed an inverse correlation with cortical thickness, with poor CCSS scores associated with cortical thinning across broad frontal areas, the right precuneus, left inferior parietal, and middle temporal regions (**Figure 1D**). CCSS also positively correlated with Sundal score (r=0.69, p<0.001, **Figure 1E**), total white matter lesion volume (r=0.70, p<0.001, **Figure 1F**), and lower corpus callosum volume (r=-0.64, p=0.001) (**Table 3**). Furthermore, CCSS was significantly associated with lower subcortical gray matter (r=-0.65, p<0.001) and cortex (r=-0.47, p=0.024) volumes, and increased ventricle volumes (r=0.48, p=0.020) (**Table 3**). These findings support the functionality of our novel CCSS tool for monitoring disease progression in CSF1R-RD patients.

### Elevated fluid biomarkers in CSF1R-RD patients

Given the critical need for identifying biomarkers that may aid in assessing severity and progression for CSF1R-RD patients, we interrogated established biomarkers of neuronal injury (NfL) ^20–22^, blood-brain barrier breaching through astrocytic damage (GFAP) ^23–25^, CSF1R ligand receptor proteins (M-CSF, IL-34) ^26–28^, and inflammation (osteopontin) ^29^, some of which may have been evaluated in smaller cohorts of CSF1R-RD patients (**Supplementary Table 1**). We measured these markers in both cerebrospinal fluid (CSF) and plasma in all subjects. In CSF, NfL levels were elevated in symptomatic *CSF1R* carriers [2972.4 (2205.2, 4646.9) pg/mL] compared to controls [489.2 (381.6, 612.75) pg/mL] (**Figure 2A**). CSF NfL levels in symptomatic *CSF1R* carriers were also elevated compared to asymptomatic carriers [505.0 (287.6, 612.9) pg/mL], but significance bordered the Bonferroni-corrected significance threshold (p<0.01). In plasma, NfL levels were significantly elevated in symptomatic *CSF1R* carriers [38.5 (14.8, 102.9) pg/mL] compared to both asymptomatic *CSF1R* carriers [6.2 (4.5, 13.1) pg/mL] and healthy controls [6.6 (4.4, 9.6) pg/mL] (**Figure 2B**); p<0.001). To assess the ability of NfL levels to distinguish between our study groups, we created receiver operator characteristic (ROC) curves. In doing so, plasma NfL [AUC = 0.88 (0.76, 1.00)] (**Figure 2C**), and to a lesser extent CSF NfL [AUC = 0.80 (0.55, 1.00)] (**Supplemental Figure 3A**), strongly differentiated between symptomatic cases and controls. In contrast, neither CSF [AUC = 0.52 (0.25, 0.78] (**Supplemental Figure 3A**) nor plasma NfL [AUC = 0.52 (0.33, 0.72)] (**Figure 2C**) levels differentiated asymptomatic from controls.

**Figure 2.**
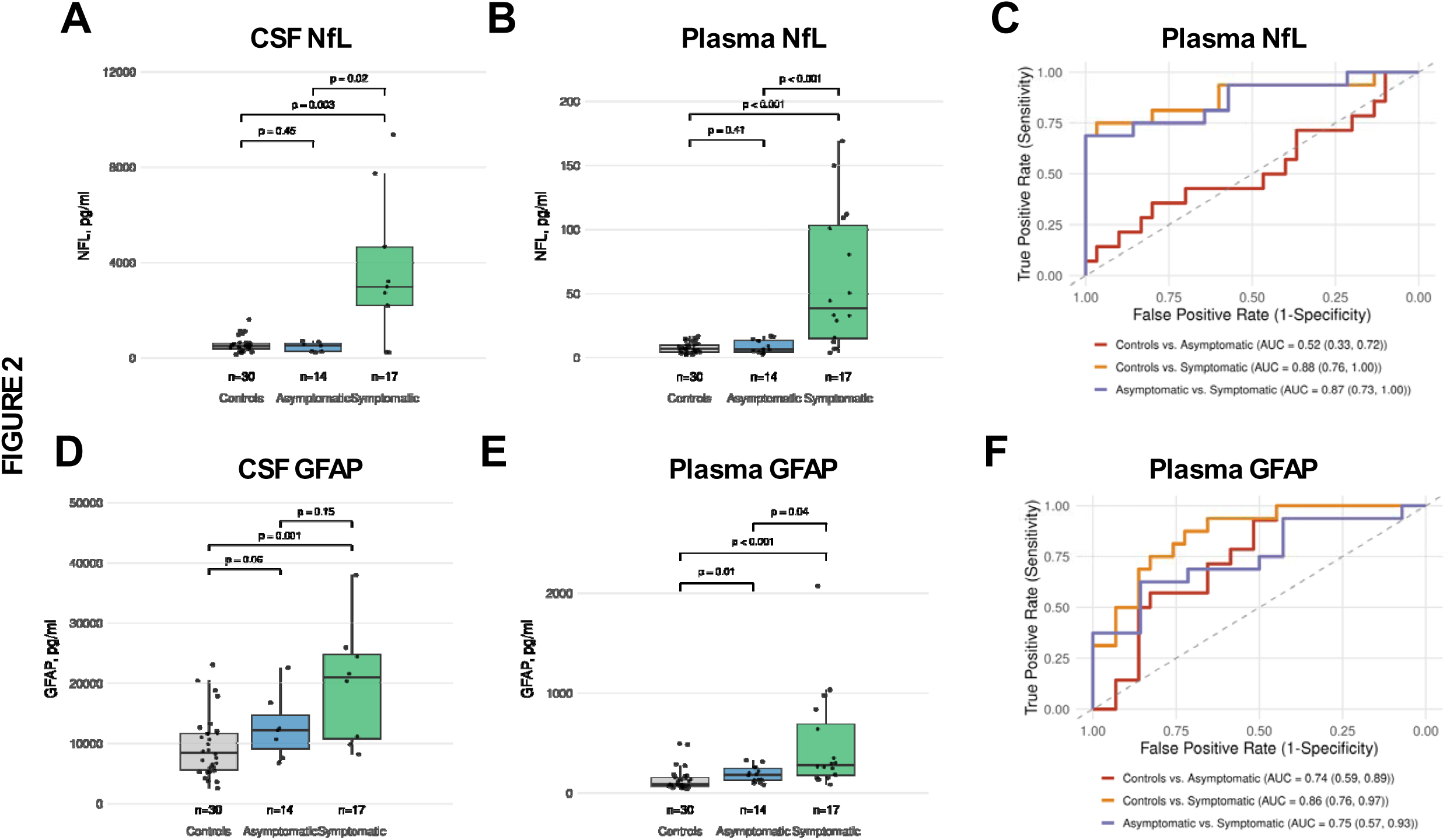
NfL and GFAP levels and discriminatory ability between symptomatic CSF1R-RD patients, asymptomatic *CSF1R* carriers, and controls. **(A-B)** NfL levels in CSF (**A**) and plasma (**B**), compared between healthy controls, asymptomatic, and symptomatic pathogenic *CSF1R* mutation carriers using Dunn’s tests (with Bonferroni-corrected significance threshold p<0.0167). (**C**) Receiver operating curves (ROC) with area under the curve (AUC) measure (with 95% confidence interval) demonstrate the ability of plasma NfL to discriminate between symptomatic, asymptomatic, and control groups. (**D-E**) GFAP levels in CSF (**D**) and plasma (**E**), compared between healthy controls, asymptomatic, and symptomatic pathogenic *CSF1R* mutation carriers using Dunn’s tests (with Bonferroni-corrected significance threshold p<0.0167). (**F**) Receiver operating curves (ROC) with area under the curve (AUC) measure (with 95% confidence interval) test the ability of plasma GFAP to discriminate between symptomatic, asymptomatic, and control groups.

Interestingly, CSF GFAP was increased in a stepwise manner from controls [8405.1 (5628.6, 11631.5) pg/mL], to asymptomatic [12102.5 (9129.2, 14654.7) pg/mL], and symptomatic [20920.3 (10805.1, 24752.7) pg/mL] *CSF1R* carriers (**Figure 2D**). Notably, the significance of increased GFAP levels was stronger in plasma, showing a clearer stepwise increase from controls [89.3 (70.5, 154.2) pg/mL], to asymptomatic [181.1 (128.9, 246.5) pg/mL], and symptomatic [280.0 (177.8, 689.4) pg/mL] *CSF1R* carriers (**Figure 2D**). Further, plasma [AUC = 0.86 (0.76, 0.97)] (**Figure 2F**), and to a lesser extent CSF [AUC = 0.83 (0.67, 0.99)] (**Supplemental Figure 3B**), significantly distinguish symptomatic *CSF1R* carriers from healthy controls. Importantly, while plasma NfL levels were not able to successfully differentiate asymptomatic *CSF1R* carriers from controls (**Figure 2C**), plasma [AUC = 0.74 (0.59, 0.89)] (**Figure 2F**), and to a lesser extent CSF [AUC = 0.70 (0.51, 0.90)] (**Supplemental Figure 3B**), GFAP levels served to more reliably distinguish between asymptomatic cases and controls.

We also measured M-CSF, IL-34, and osteopontin in CSF and plasma. M-CSF levels were elevated in symptomatic patients [CSF: 13.3 (10.8, 15.8), plasma: 31.4 (26.4, 35.2), pg/mL] compared to controls [CSF: 5.5 (4.2, 6.7), plasma: 23.8 (21.0, 27.4), pg/mL] (**Supplementary Figures 4A-B**), as well as when comparing asymptomatic individuals [CSF: 7.9 (6.8, 10.4), plasma: 29.8 (26.4, 36.3) pg/mL] to controls [CSF: 5.5 (4.2, 6.7), plasma: 23.8 (21.0, 27.4) pg/mL] (**Supplementary Figures 4A-B**). Similar findings were observed for IL-34, where significantly higher levels were observed in symptomatic [CSF: 132.3 (95.2, 224.4), plasma: 2.5 (2.0, 3.2), pg/mL] and asymptomatic [CSF: 142.5 (123.6, 151.6), plasma: 2.7 (1.7, 3.1), pg/mL] individuals compared to controls [CSF: 58.2 (46.1, 71.7), plasma: 1.5 (1.1, 1.9), pg/mL] (**Supplementary Figures 4C-D**). However, there were no differences in either M-CSF or IL-34 levels between symptomatic and asymptomatic individuals (**Supplementary Figure 4A-D**).

Furthermore, osteopontin levels did not differ significantly between symptomatic, asymptomatic, and healthy control groups in either CSF or plasma (**Supplementary Figures 4E-F)**. Overall, our data suggests that plasma GFAP may be more sensitive to early pathological changes preceding symptom onset, while plasma NfL better reflects disease progression after initial symptom onset.

### GFAP and NfL biomarkers may inform on clinical and neuroanatomical changes in CSF1R-RD patients

To determine if fluid biomarkers inform clinical assessments, we performed associations between NfL and GFAP with MoCA and CCSS. NfL demonstrated the most robust correlations with CCSS in both plasma (r=0.76, p<0.001) (**Figure 3A**) and CSF (r=0.74, p=0.002) (**Table 3**), but also with MoCA to a lesser degree [plasma: (r=-0.61, p<0.001), CSF: (r=-0.64, p=0.010)] (**Table 3**). Increased GFAP levels were also highly associated with worsened CCSS especially in plasma (r=0.67, p<0.001) (**Figure 3B**), but also in CSF (r=0.63, p=0.011) (**Table 3**). Plasma GFAP (r=-0.49, p=0.007), but not CSF GFAP (r=0.63, p=0.011), also associated with MoCA scores (**Table 3**). Weaker to no correlations were observed between other fluid biomarkers (M-CSF, IL-34, osteopontin) and either MoCA or CCSS scores (**Table 3**).

**Figure 3.**
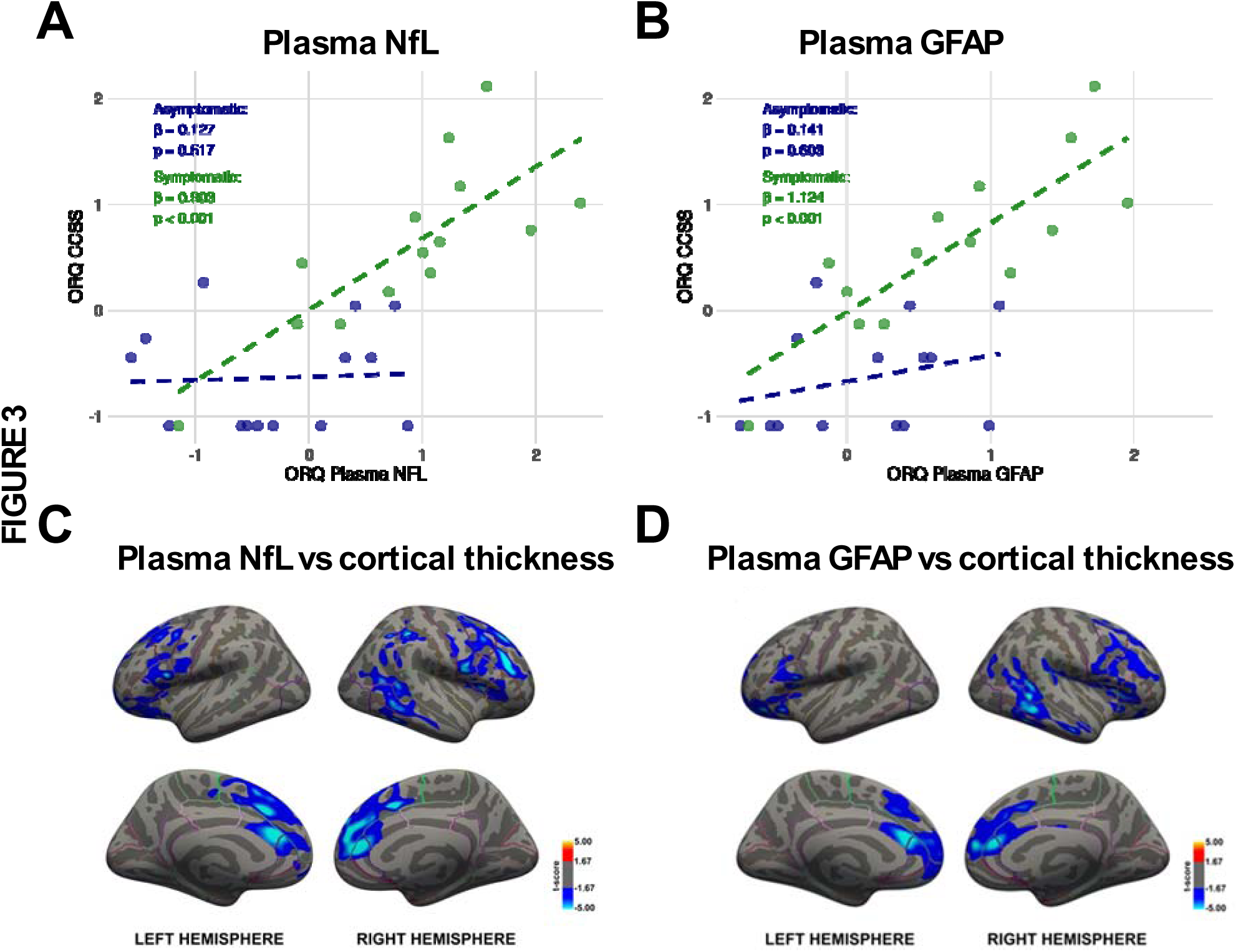
Plasma NfL and GFAP sensitively reflect CCSS and physiological changes associated with CSF1R-RD. **(A-B)** Scatterplots of ranked relationships (ORQ-transformed) between CSF1R-RD Clinical Severity Score (CCSS) and plasma NfL (**A**) and GFAP (**B**) indicate a strong positive relationship between these biomarkers and CCSS in symptomatic patients. (**C-D**) Correlations of plasma NfL (**C**) and plasma GFAP (**D**) with cortical thickness measured with 3D T1-weighted MPRAGE scans across multiple brain regions indicate that both biomarkers are reflective of the physiological changes associated with CSF1R-RD disease progression.

Next, we evaluated the relationships between the levels of fluid biomarkers with cortical thickness measurements from 3D T1-weighted MPRAGE scans. Interestingly, plasma NfL was inversely correlated with cortical thickness across broad prefrontal regions, cingulum, precentral gyrus, as well as the right inferior parietal lobe and lateral temporal lobe (**Figure 3C**).

Additionally, plasma NfL correlated strongly with Sundal score (r=0.71, p<0.001), higher total white matter lesion volume (r=0.56, p=0.005), and lower normalized brain volumes (r=-0.70, p<0.001) (**Table 4**). CSF NfL showed even stronger correlations with Sundal score (r=0.85, p<0.001) and total white matter lesion volume (r=0.79, p<0.001), while showing a trend toward correlation with normalized brain volume (r=-0.66, p=0.012), but not meeting the Bonferroni-corrected significance threshold of p<0.01 (**Table 4**).

**Table 4.** Testing the correlation between fluid biomarkers and neuroanatomical changes.

|  | Plasma |  |  |  |  | CSF |  |  |  |  |
| --- | --- | --- | --- | --- | --- | --- | --- | --- | --- | --- |
|  | M-CSF | IL 34 | NfL | Osteopontin | GFAP | M-CSF | IL 34 | NfL | Osteopontin | GFAP |
|  | Spearman's r, p-value |  |  |  |  | Spearman's r, p-value |  |  |  |  |
| **Sundal score** | -0.22,<br>0.308 | 0.18,<br>0.405 | **0.71,**<br>**<0.001** | 0.04,<br>0.859 | 0.36,<br>0.088 | 0.40,<br>0.153 | **-0.61,**<br>**0.04** | **0.85,**<br>**<0.001** | 0.59,<br>0.058 | **0.71,**<br>**0.004** |
| **Sundal score (WM)** | -0.22,<br>0.308 | 0.24,<br>0.250 | **0.69,**<br>**<0.001** | 0.09,<br>0.670 | 0.36,<br>0.087 | 0.43,<br>0.125 | **-0.64,**<br>**0.035** | **0.86**<br>**<0.001** | **0.71,**<br>**0.014** | **0.74,**<br>**0.003** |
| **Sundal score (atrophy)** | -0.08,<br>0.694 | 0.06,<br>0.790 | **0.72,**<br>**<0.001** | -0.06,<br>0.781 | 0.40,<br>0.056 | 0.51,<br>0.064 | -0.29,<br>0.380 | **0.71,**<br>**0.004** | 0.39,<br>0.241 | **0.64,**<br>**0.013** |
| **Total WM Lesion Volume (mm<sup>3</sup>)** | -0.15<br>0.496 | -0.02<br>0.925 | **0.56**<br>**0.005** | -0.15<br>0.512 | 0.31,<br>0.156 | 0.30,<br>0.293 | **-0.64,**<br>**0.033** | **0.79,**<br>**<0.001** | **0.69,**<br>**0.023** | **0.65,**<br>**0.012** |
| **Normalized Brain Volume** | 0.14,<br>0.530 | 0.02,<br>0.938 | **-0.70,**<br>**<0.001** | 0.03,<br>0.889 | **-0.44,**<br>**0.038** | -0.49,<br>0.081 | 0.51,<br>0.114 | **-0.66,**<br>**0.012** | -0.36,<br>0.273 | -0.53,<br>0.057 |
| **Normalized WM Volume** | 0.10,<br>0.655 | -0.10,<br>0.642 | **-0.57,**<br>**0.005** | 0.29,<br>0.188 | -0.32,<br>0.134 | -0.38,<br>0.177 | -0.06,<br>0.863 | -0.37,<br>0.193 | -0.32,<br>0.339 | -0.40,<br>0.156 |
| **Corpus Callosum Total Volume (mm<sup>3</sup>)** | -0.06,<br>0.771 | -0.31,<br>0.154 | **-0.55,**<br>**0.007** | 0.10,<br>0.650 | -0.33,<br>0.130 | -0.33,<br>0.253 | 0.15,<br>0.673 | -0.40,<br>0.160 | -0.20,<br>0.558 | -0.35,<br>0.221 |
| **Cortex Volume (mm<sup>3</sup>)** | -0.32,<br>0.142 | -0.11,<br>0.630 | **-0.56,**<br>**0.006** | -0.24,<br>0.288 | -0.32,<br>0.136 | -0.38,<br>0.186 | -0.07,<br>0.839 | -0.27,<br>0.349 | -0.26,<br>0.435 | -0.20,<br>0.483 |
| **Cerebral WM Volume (mm<sup>3</sup>)** | -0.32,<br>0.133 | -0.24,<br>0.261 | **-0.61,**<br>**0.002** | -0.11,<br>0.639 | -0.27,<br>0.206 | -0.50,<br>0.072 | -0.26,<br>0.435 | -0.16,<br>0.584 | -0.19,<br>0.576 | -0.31,<br>0.288 |
| **Subcortical Gray Matter Volume (mm<sup>3</sup>)** | -0.23,<br>0.293 | -0.18,<br>0.400 | **-0.80,**<br>**<0.001** | -0.16,<br>0.466 | **-0.49,**<br>**0.019** | **-0.58,**<br>**0.032** | -0.05,<br>0.882 | -0.46,<br>0.101 | -0.37,<br>0.261 | -0.53,<br>0.057 |
| **Ventricles Volume (mm<sup>3</sup>)** | -0.23,<br>0.287 | -0.29,<br>0.178 | **0.62,**<br>**0.002** | -0.12,<br>0.593 | **0.43,**<br>**0.041** | 0.50,<br>0.069 | -0.59,<br>0.061 | **0.80,**<br>**<0.001** | 0.31,<br>0.356 | **0.67,**<br>**0.011** |

Spearman's test of correlation selected for untransformed, non-normal data. After applying a Bonferroni correction for multiple testing, p-values < 0.01 were considered significant. CSF -cerebrospinal fluid, M-CSF - macrophage colony stimulating factor, IL-34 - interleukin-34, NfL-neurofilaments light chains, GFAP - glial fibrillary acidic protein, WM -white matter.

Plasma GFAP levels were also inversely correlated with cortical thickness in the bilateral frontal operculum, cingulum, superior frontal gyrus, as well as the right inferior parietal and lateral temporal lobes (**Figure 3D**). Plasma GFAP showed a trend toward correlation with Sundal score (r=0.36, p=0.088) and smaller normalized brain volumes (r=-0.44, p=0.038), but did not reach significance with total white matter lesion volume (r=0.31, p=0.156) (**Table 4**). CSF GFAP also demonstrated strong correlations with Sundal score (r=0.71, p=0.004), but showed only a trend toward correlation with total white matter lesion volume (r=0.65, p=0.012) and normalized brain volume (r=-0.53, p=0.057) (**Table 4**). Of note, the levels of plasma or CSF for M-CSF, IL-34, and osteopontin exhibited weak to no associations with Sundal scores or white matter lesions (**Table 4**). Overall, plasma NfL and GFAP showed the strongest associations with CCSS and MoCA clinical assessments, as well as with neuroanatomical changes including cortical thinning, normalized brain volumes, Sundal scoring, and total white matter lesions. Taken together, these findings provide strong evidence to support the utility of plasma NfL and GFAP as biomarkers for tracking CSF1R-RD disease progression, with plasma GFAP displaying the additional capability of identifying the earliest stages of disease.

## DISCUSSION

Our study leveraged a well-characterized cohort of 31 individuals carrying a pathogenic *CSF1R* variant, representing, to our knowledge, the largest cohort of its kind to date. All individuals underwent fluid biomarker testing, with 23 undergoing additional neuroimaging biomarker testing, further enriching our dataset. Importantly, none of the participants received therapeutic interventions, positioning this cohort as a valuable untreated reference point for evaluating clinical trials in CSF1R-RD. Additionally, we developed a novel clinical diagnostic tool, the CSF1R-RD Clinical Severity Score (CCSS), which we believe is an accurate, reproducible, and sensitive scale for monitoring symptoms in patients with CSF1R-RD. This study aimed to identify both fluid and neuroimaging biomarkers for CSF1R-RD that can inform the optimal timing of treatment administration to maximize therapeutic benefit, while also providing sensitive quantitative measurements to monitor disease progression.

Neuroimaging biomarkers have been extensively studied in CSF1R-RD research, with some accurately reflecting established pathological features of the disease. However, none have demonstrated sufficient reliability in identifying the critical transition from presymptomatic to symptomatic stages. Further, the sensitivity of neuroimaging biomarkers to capturing nuanced changes in disease progression remains uncertain and continues to be a subject of debate.

Another important limitation to the utility of neuroimaging biomarkers in CSF1R-RD is the significant overlap in imaging features with other leukoencephalopathies, including AARS-related leukoencephalopathy and CSF1R/AARS-negative cases ^17,19^. In our study, symptomatic patients had significantly higher Sundal scores and were characterized by more severe white matter lesions and brain atrophy measured by Sundal subscales. Interestingly, brain volume or corpus callosum volume could not significantly discriminate symptomatic from asymptomatic individuals in our cohort, which may be related to the small sample size. White matter lesions were highly elevated in symptomatic patients compared with asymptomatic patients, suggesting that white matter involvement in CSF1R-RD pathology may be a marker for conversion to the symptomatic phase. In our cohort, we also found a significant correlation between the severity of neuroimaging changes (as measured by the Sundal scale and brain and white matter lesion volumes) and the clinical symptoms of the disease, as assessed by the CCSS and MoCA. Notably, we found that reduced cortical thickness, most prominently across the prefrontal cortex, is significantly correlated with poor clinical status. This result supports work by Kinoshito et al. ^30^, which demonstrated that MR images were associated with more advanced pathological lesion stages. As demonstrated in this paper, the Sundal scale ^19^ has strong potential for describing the clinical status in CSF1R-RD; however, its semi-quantitative nature may limit its utility for detecting subtle changes in longitudinal cohorts, in which case radiologic assessment based on volume measurements is likely to be superior. It is worth noting that these changes may already be reflected in increases in fluid biomarkers. When assessing asymptomatic patients, it is also important to consider that white matter changes may occur during the preclinical period of CSF1R-RD or may be attributed to other causes. Further studies, especially those conducted in longitudinal cohorts using precise radiological parameter measurements, are needed to determine the usefulness of MRI in monitoring the progression of CSF1R-RD.

Fluid biomarkers offer significant advantages over neuroimaging biomarkers for tracking CSF1R-RD progression and detecting early symptom onset. They offer a high degree of reliability, reproducibility, and can be collected through minimally invasive procedures, making them far more practical for routine clinical use. Additionally, fluid biomarkers enable more frequent and accessible testing, which is critical for capturing more subtle changes in disease progression and creating an optimized care plan for CSF1R-RD patients. So far, basic CSF analyses (cell count, glucose and protein levels; presence of inflammatory cells; oligoclonal bands) have proven to be unremarkable in the monitoring of CSF1R-RD ^31^. Consequently, there remains an urgent need to identify alternative fluid biomarkers that can accurately reflect CSF1R-RD disease progression.

NfL is well-established as a robust biomarker for other rapidly progressive neurodegenerative diseases, but it is known to lack disease-specificity ^22, 32,20^. Previous work by Hayer et al. explored the utility of NfL as a robust fluid biomarker for CSF1R-RD and found elevated levels of NfL in both CSF and plasma in symptomatic patients ^33^. Notably, they showed that NfL can differentiate CSF1R-RD from multiple sclerosis (MS), a common misdiagnosis of CSF1R-RD. In the Hayer et al. study, MS was characterized by significantly lower NfL levels, which may indicate a faster progression of neurodegeneration in CSF1R-RD. The first clinical symptoms may occur with a delay in relation to the pathological process, and an increase in NfL in asymptomatic or mildly symptomatic patients may indicate the initiation of the pathogenic process. This finding has profound implications for currently available treatments, such as off-label HSCT. The central nervous system is accessible only when the blood-brain barrier is disrupted. Thus, patients with sufficiently advanced disease, in which the blood-brain barrier has been disrupted, will be most likely to benefit from treatment. Available data supports offering treatments only after the emergence of symptomatic disease ^13^. Identifying this moment based on objective biomarkers could allow for optimization of therapy efficacy.

In our work, we also found a significant correlation between clinical severity diagnostics (both CCSS and MoCA) and NfL levels, both in serum and CSF. Interestingly, in the study by Serreno et al. ^34^, the NfL level correlated with the degree of spasticity, fatigue, depression and MoCA; however, it is worth noting that our study was conducted on a larger cohort, which may strengthen the analysis. It is also important to note that previous studies ^34, 35^ relied on scales addressing only specific aspects of the CSF1R-RD clinical presentation. In contrast, the use of our CCSS allowed a more comprehensive assessment of the disease overall clinical symptoms. Based on our findings, NfL has a promising potential as a robust biomarker of CSF1R-RD disease progression and could be used to track treatment efficacy in clinical trials. NfL is not able to differentiate CSF1R-RD cases from other rapidly neurodegenerative diseases due to its non-specific behavior and should be used only in patients with a genetic diagnosis of CSF1R-RD. It should be noted that NfL is not typically substantially elevated in the early stages of many different glial-cell-related conditions and may not be sensitive to the initial phase of axonal damage ^20–22, 36^. Our study confirms the utility of NfL in a larger patient cohort, further reinforcing its strong association with clinical symptom severity and structural CNS pathology. Notably, this association was supported by a novel integration of neuroimaging analysis, adding strength and specificity to our findings

For leukoencephalopathies that are primarily characterized by cellular dysfunction at the glial cell level, GFAP may better reflect the pathological process, especially in early disease stages. GFAP has already proven to be a particularly robust biomarker for X-linked adrenoleukodystrophy, metachromatic leukodystrophy, and Alexander disease ^24, 25, 37^. Despite its established relevance in other conditions, this biomarker has not been previously evaluated in the context of CSF1R-RD. Our study provides the first data supporting its potential utility in clinical assessment of this disease. In our cohort, plasma GFAP was notably elevated in asymptomatic *CSF1R* pathogenic variant carriers when compared to both healthy controls and symptomatic CSF1R-RD patients. This finding suggests that GFAP may be a robust biomarker for detecting early symptom onset, particularly compensating for NfL’s known limitations in detection at this stage of disease. CSF GFAP was weakly correlated with the CCSS but did not show a correlation with MoCA, most likely due to small sample size.

Based on the strong and consistent association observed between NfL levels, clinical symptoms, and neuroimaging biomarkers in our cohort, as well as supporting evidence from previous studies (*35,36*), we propose a complementary role for NfL and GFAP in the clinical management of CSF1R-RD. While GFAP demonstrates exceptional sensitivity in distinguishing early-stage patients from healthy controls and may help identify the optimal timing for therapeutic interventions, NfL appears to be more effective for tracking disease progression following the onset of symptoms.

This is also the first study to evaluate potential disease-specific biomarkers that are directly implicated in the pathophysiology of CSF1R-RD. IL-34 is a ligand for CSF1R, therefore directly implicating in pathophysiological processes related to CSF1R-RD ^38^. Therefore IL-34 could serve as a more specific marker for CSF1R. Research on IL-34 activation in rodent microglia and human macrophages suggests that it has distinct properties compared to colony-stimulating factor 1 (CSF1), resulting in an anti-inflammatory and reparative phenotype. However, chronic IL-34 activation of microglia enhances the expression of pro-inflammatory cytokines, contributing to neuroinflammation during the later stages of neurological disorders ^26, 27, 38^. In our cohort, IL-34 levels were elevated in all patients with pathological *CSF1R* variants compared with healthy controls, suggesting a potential role for IL-34 in the preclinical phase of CSF1R-RD. However, it remains uncertain whether IL-34 levels can reliably detect the clinical onset of CSF1R-RD. In our cohort, higher IL-34 levels were observed in patients with less extensive white matter lesions, suggesting that IL-34 may have a protective role in the preclinical stage.

Longitudinal studies tracking changes in IL-34 levels could potentially identify the onset of the pathophysiological process earlier than other biomarkers. However, this hypothesis cannot be confirmed based on the current study. Another limitation of IL-34 is its lack of correlation with CSF1R-RD symptoms, which may point to the involvement of compensatory mechanisms in the preclinical phase of the disease.

Another potential biomarker for CSF1R-RD is M-CSF, which signals through its receptor, CSF-1R, and thus, like IL-34, is involved in the pathophysiology of CSF1R-RD. Interestingly, mice deficient in *CSF1R* expression exhibit a more pronounced phenotype than those lacking in M-CSF ^28^. In our cohort, M-CSF, like IL-34, was elevated in both asymptomatic and symptomatic carriers of pathogenic *CSF1R* variants compared with healthy controls and did not correlate with clinical status. However, the utility of M-CSF as a marker for the pathophysiological processes underlying CSF1R-RD requires further investigation.

In our study, we utilized a robust cohort consisting of 31 individuals with a pathogenic CSF1R variant, which also included fluid biomarker and clinical symptom data—representing, to our knowledge, the largest cohort of its kind to date. Twenty-three of these individuals also underwent neuroimaging biomarker testing, further enhancing the depth of our analysis.

Another strength of our study is that none of the individuals included underwent therapeutic interventions. These individuals illustrate the natural disease course, and can serve as an untreated reference point for evaluating clinical trials in CSF1R-RD. However, our studies also present some limitations. Not all patients in our cohort underwent CSF biomarker testing. Therefore, the strength of CSF biomarker results may have been limited. Furthermore, this cohort lacks follow-up data, making it unclear how useful these biomarkers would be in longitudinal cohorts. Further studies are needed to evaluate the utility of these biomarkers in monitoring treatment response and to confirm their clinical applicability in therapeutic settings. Another limitation is the lack of evaluation of CLP, which has been identified as a strong CSF1R-RD biomarker in previous studies.^34, 35^

Ultimately, our study demonstrates that the plasma biomarkers NfL and GFAP are promising tools for detecting the onset and progression of CSF1R-RD, providing a robust means to evaluate patients and serving as a valuable reference for future clinical trials.

## MATERIALS AND METHODS

### Study design

Our observational study was comprised of 31 individuals, 11 males (35.5%) and 20 females (64.5%), with mean age 47.9±15.9 years, ranging from 20 to 77 years of age, 11 males (35.5%) and 20 females (64.5) carrying pathogenic variants of *CSF1R*, including 17 symptomatic patients with mean age of onset 49.4±14.8 years of age with mean age of symptoms’ onset 43.4±15.6 years and 14 asymptomatic individuals with mean age 46.1±17.5 years and 30 controls, 12 females (40%) and 18 males (60%), with mean age 51.4±13.0 years. Cohort characteristics are summarized in **Table 1**. All asymptomatic and symptomatic patients were heterozygous *CSF1R* variant carriers. A list of *CSF1R* variants from this cohort are listed in **Supplementary Figure 1**. Informed consent was obtained from all individuals according to standard protocols approved by the Mayo Clinic Institutional Review Board (IRB) ethics committee. The study protocol was approved by the Mayo Clinic IRB. Outliers were identified by examining individual datapoints that fell above or below the 1st or 99th quantile for each biomarker measurement and imputed with NA if clinician deemed likely the result of a technical error.

### Neurological assessment

Every participant underwent a structured neurological assessment. We developed a semi-quantitative scale that captures all possible symptoms of CSF1R-RD (henceforth referred to as CSF1R Clinical Severity Score (CCSS)). It assesses the patient in five domains: cognition, mood and affect, cranial nerves, motor function and sensory function. For each symptom assessed, the patient was given points according to the severity of the symptom rated as mild (1 point), moderate (2 points) or severe (3 points), then the points were counted in each category and summed into a total CCSS. It is important to note that our CCSS is sensitive enough to detect even minor cognitive and motor deviations-therefore, it is possible that a patient who is classified as asymptomatic may still have points on this scale. Symptomatic CSF1R-RD patients were defined as those exhibiting clear, progressive signs or symptoms of CSF1R-RD and having a CCSS greater than 5. Patients with a CCSS of 5 or less were considered asymptomatic. The detailed scoring system can be found in **Supplementary Tool 1**. Cognitive functions were also assessed with The Montreal Cognitive Assessment (MoCA). CCSS and MoCA were then correlated with neuroimaging results and fluid biomarkers. We analyzed the total CCSS to better reflect biomarker results with the overall clinical status of CSF1R-RD patients. All research subjects both symptomatic and asymptomatic were examined by the same neurologist (ZKW).

### Neuroimaging assessment

Twenty-three participants with *CSF1R* pathogenic variants (13 asymptomatic and 10 symptomatic) had 3-Tesla MRI of the brain available, obtained within one week of biomarker collection. Scans were analyzed by a single neuroradiologist (EHM). Sequences included in this analysis consisted of a 3D T1-weighted MPRAGE and 3D T2 FLAIR. All scans were graded using the Sundal scale ^19^, including total, white matter lesion, and atrophy scores with the reader blinded to the clinical information. White matter lesion volume was calculated from the 3D FLAIR using a deep-learning based approach implemented in Lesion Segmentation Tool (LST-AI; https://github.com/CompImg/LST-AI) ^39^ with subsequent manual verification of segmentations.

Cortical thickness data were derived for each patient from the 3D T1-weighted MPRAGE images using the default “recon-all” pipeline implemented in FreeSurfer 7.2 (http://surfer.nmr.mgh.harvard.edu). The thickness values were entered into a general linear model (mri_glmfit) for each of the primary variables of interest, including CCSS, MoCA, plasma GFAP, and plasma NfL levels. Age and sex were also entered as covariates. Permutation-based, clusterwise-corrected inference was then performed using mri_glmfit-sim (version 7.4.1) with 10,000 permutations and a clusterwise p-value threshold (cwp) of 0.05. Significant clusters were identified and characterized with MRI_surfcluster, while mri_segstats provided summary statistics for each labeled cluster. FreeSurfer segmentations were also used to derive normalized brain volume (total brain volume normalized by total intracranial volume) and corpus callosum volume.

### Biomarker assessments

Plasma was collected from all patients and controls involved in this study, and CSF samples were collected from 16 individuals carrying pathogenic *CSF1R* variants and from all controls. Neurofilament light chains (NfL), macrophage colony stimulation factor (M-CSF), Glial fibrillary acidic protein (GFAP), interleukin-34 (IL-34) and osteopontin were assessed in both plasma and CSF.

All analytes were measured using the Simoa® HD-1 analyzer and commercially validated Simoa® HD-1 kits, Human Neurology 2-Plex assay for GFAP and NfL (Quanterix, Billerica, MA, USA). The plasma and CSF samples were assessed using 96-well plates. Each measure was repeated twice per Simoa® platform guidelines, and the mean of both measurements was provided along with the coefficient of variation (CV).

Outliers were identified by examining individual datapoints that fell above or below the 1st or 99th quantile for each biomarker measurement and removed from subsequent analyses if clinician deemed likely the result of a technical error.

### Statistical analysis

Descriptive statistics, including mean, standard deviation, median, and interquartile range (IQR), were calculated using the ‘stats’ package in R ^40^. Differences in fluid biomarker levels across symptomatic, asymptomatic, and healthy control groups were assessed using the Kruskal-Wallis test with accompanying post-hoc Dunn’s tests. Differences in neuroimaging levels across asymptomatic and symptomatic groups were assessed using the Mann-Whitney U test. Spearman’s rank correlations were used to examine the relationships between fluid biomarkers, neuroimaging biomarkers, CCSS, and MoCA. To further visualize the relationships captured in the rank-based Spearman’s correlations, CCSS, MoCA, Total WM Lesion Volume, Normalized Brain Volume,plasma NfL, and plasma GFAP were transformed using rank-ordered quantile normalization (ORQ) and visualized in scatterplots, with linear regression models used to fit lines and test linear associations between the ranks of biomarker and CCSS, with an interaction term for patient status ^41^. Receiver-operator characteristic curves with area under the curve (AUC) scores were generated using the ‘pROC’ package in R to compare the discriminatory ability of biomarkers of interest for patient status comparison groups ^42^. The level of statistical significance was set at p-value < 0.05, and all tests were two-tailed. Where applicable, tests were accompanied by Bonferroni-adjusted p-value significance thresholds to account for multiple comparisons. All visualizations were generated using the ‘ggplot2’ package in R ^43^.

## Acknowledgments

We are grateful to all participants and their caregivers.

## Author contributions

Conceptualization: SG, GP, LP, MP, ZKW

Methodology: TC, MR, EHM, MP

Investigation: TC, MR, EHM, KJW, JD, YS, AJS, SG, GP, MBJ, RCS

Visualization: TC, MR, EHM, MP

Funding acquisition: SG, GP, LP, MP, ZKW

Project administration: LP, MP, ZKW

Supervision: LP, MP, ZKW

Writing – original draft: TC, MR, EHM, MP

Writing – review & editing: TC, MR, KJW, JD, YS, AJS, SG, GP, MBJ, RSC, EHM, LP, MP, ZKW

## Data and materials availability

All data supporting the findings of this study are available within the paper and its supplementary materials. Due to patient privacy concerns and institutional regulations, some individual-level clinical data cannot be publicly shared. Access to this data may be granted upon reasonable request and will require a completed Materials Transfer Agreement (MTA) and approval from the corresponding institutional review board. Requests for data and materials should be directed to the corresponding author

