## Supplementary material for "Fluid and Neuroimaging Biomarkers in Microgliopathy Colony-Stimulating Factor-1 Receptor-Related Disorders": Supplementary Materials.docx

**Suplementary Figure 1.**

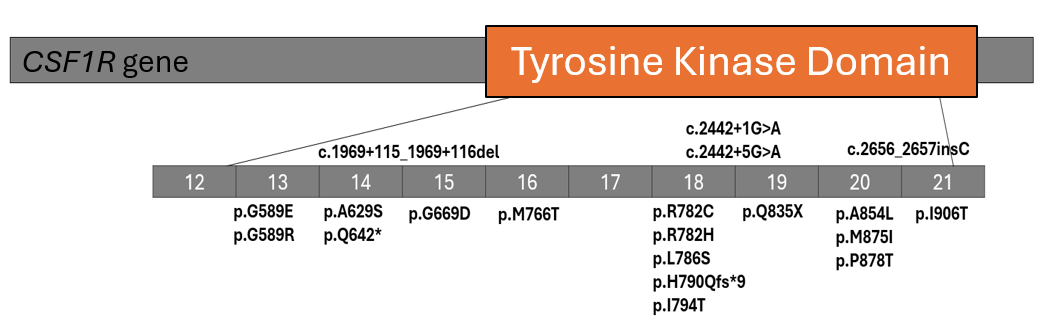

| **No.** | ***CSF1R* variant** |
| --- | --- |
| **1** | p.G589E |
| **2** | p.A629S |
| **3** | p.Q642* |
| **4** | p.G669D |
| **5** | c.1969+115_1969_116del |
| **6** | p.M766T |
| **7** | c.2442+1G>A |
| **8** | p.R782H |
| **9** | p.L786S |
| **10** | p.H790Qfs*9 |
| **11** | p.I794T |
| **12** | p.P878T |
| **13-14** | c.2656_2657insC |
| **15** | p.G589R |
| **16-17** | p.G589E |
| **18** | p.A629S |
| **19-20** | c.1969+115_1969_116del |
| **21** | p.I794T |
| **22** | c.2442+1G>A |
| **23** | c.2442+5G>A |
| **24** | p.M875I |
| **25** | p.R782C |
| **26** | p.Q835X |
| **27** | p.A854L |
| **28** | p.P878T |
| **29-31** | c.2656_2657insC |

**Supplementary Figure 1.** ***CSF1R* varaints detected in the CSF1R-RD patient cohort (related to Table 1).** Schematic represenation (top) of the location of the different *CSF1R* mutations (listed at the bottom) within the tyrosine kinase domain. Note cases 1-14 are asymptomatic and 15-31 are symptomatic *CSF1R* carriers.

**Supplementary Figure 2.**

**Supplementary Figure 2. Neuroimaging markers in asymptomatic and symptomatic *CSF1R* carriers (related to Figure 1).** Sundal – atrophy (**A**), Sundal – white matter (**B**), Normalized brain volume (**C**), Normalized white matter volume (**D**), Corpus callosum total volume (**E**), Cerebral white matter volume (**F**), Subcortical grey matter volume (**H**), Ventricle volume **(I**), and Cortex volume (**J**), levels compared between symptomatic and asymptomatic groups using Mann Whitney U Tests.

**Supplementary Figure 3.**

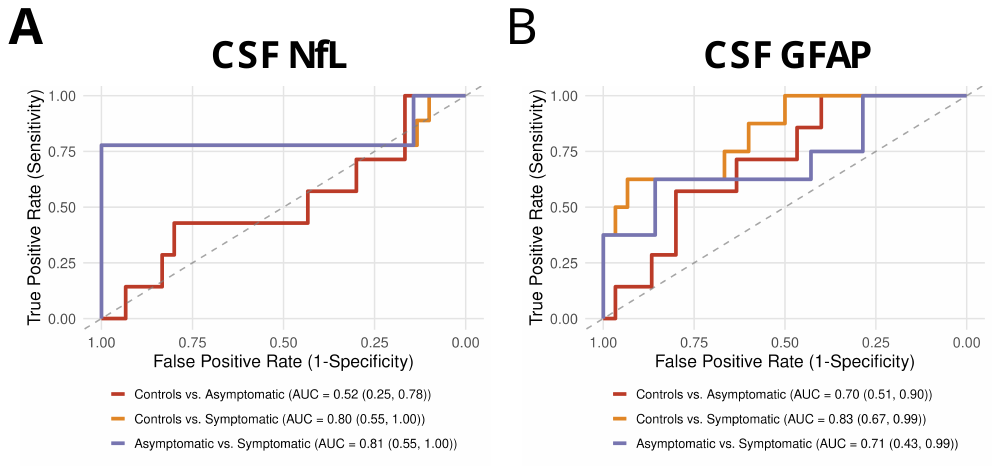

**Supplementary Figure 3. The ability of CSF NfL and GFAP to discriminate between symptomatic *CSF1R* patients, asymptomatic *CSF1R* carriers, and controls (related to Figure 2).** Receiver operating curves (ROC) with area under the curve (AUC) measures (with 95% confidence interval) test for CSF NfL (**A**) and CSF GFAP (**B**).

**Supplementary Figure 4.**

**Supplementary Figure 4.** **M-CSF, IL-34 and osteopontin levels in CSF and plasma of CSF1R-RD cases and controls (related to Figure 2).** M-CSF CSF (**A**) and plasma (**B**), IL-34 CSF (**C**) and plasma (**D**), and osteopontin CSF (**E**) and plasma (**D**) levels compared between symptomatic, asymptomatic, and control groups using Dunn’s tests. P-values < 0.0167 are considered significant after applying a Bonferroni correction for multiple testing.

**Supplementary Table 1. Summary of studies evaluating NfL, GFAP, M-CSF, IL-34, osteopontin, and other biomarkers in CSF1R-RD cases.**

|  | **This report** | **Hayer et al. (2018) (1)** | **Hayer et al.**  **(2022) (*7*)** | **Serreno et al.**  **(2024) (3)** |
| --- | --- | --- | --- | --- |
| No. symptomatic CSF1R-RD (n) | 17 | 10 | 14 | 11 |
| No. asymptomatic CSF1R carriers | 14 | 7 | 7 | 7 |
| No. controls | 30 | 26 | 10 | 15 |
| Studied biomarkers: | NfL  GFAP  M-CSF  IL-34  Osteopontin | NfL | CHIT | NfL  CHIT  CHI3L2 |
| Elevated in symptomatic cases (vs controls) | NfL*  GFAP***  M-CSF*  IL-34* | NfL* | CHIT* | NfL*  CHIT*  CHI3L2* |
| Elevated in asymptomatic cases (vs controls) | GFAP*  IL-34*  M-CSF* | NfL* | CHIT not elevated | CHIT  NfL |
| Correlation with clinical symptoms | NfL* (with CCSS and MoCA)  GFAP*** (with CCSS and MoCA,) | Not done | CHIT  No correlation with MoCA and Barthel Index  Negative correlation with disease duration** | CHIT and  CHI3L2 correlation with: SPRS, PHQ-8, GAD-7, FSS, MoCA, PSS, MCS, SF-12  NfL correlation with SPRS, FSS, PHQ-8, PCS, MCS |
| *Both in plasma and CSF  **Only CSF  ***Only plasma | | | | |

Summary of studies on biomarkers in CSF1R-RD. NfL- neurofilaments light chains, glial fibrillary acidic protein (GFAP), macrophage colony stimulating factor (M-CSF), interleukin-34 (IL-34), CHIT – Chitotriosidase, CHI3L2 - chitinase 3-like 2. CCSS – Colony stimulation factor-1 Receptor Related Disorder Clinical Severity Scale. *Statistically significant only in plasma. SPRS - Spastic Paraplegia Rating Scale, PHQ-8 - Patient Health Questionnaire-8, GAD-7 - Generalized Anxiety Disorder 7-item, FSS- Fatigue Severity Scale, PSS - Perceived Stress Scale, MCS - Malnutrition Care Score, SF-12 - 12-Item Short Form Survey
