## Supplementary material for "Fluid and Neuroimaging Biomarkers in Microgliopathy Colony-Stimulating Factor-1 Receptor-Related Disorders": Supplementary Tool 1.docx

**CSF1R-RD Clinical Severity Score (CCSS)**

| **Domain I: Cognition** | | | | |
| --- | --- | --- | --- | --- |
|  | Normal (0) | Mild (1) | Moderate (2) | Severe (3) |
| Working memory |  |  |  |  |
| Episodic memory |  |  |  |  |
| Remote memory |  |  |  |  |
| **Memory total (0-9 points):** |  | | | |
| Spontaneous speech |  |  |  |  |
| Naming |  |  |  |  |
| Repetition |  |  |  |  |
| Reading |  |  |  |  |
| Comprehension |  |  |  |  |
| **Language total (0-15 points)** |  | | | |
| Aphasia |  |  |  |  |
| Agraphia |  |  |  |  |
| Acalculia |  |  |  |  |
| Apraxia |  |  |  |  |
| Attention |  |  |  |  |
| **Higher cortical functions total (0-15 points)** |  | | | |
| **Cognition total (0-39 points)**  **Language + Memory + Higher Cortical Function** |  | | | |

| **Domain II: Mood and Affect** | | | | |
| --- | --- | --- | --- | --- |
|  | Normal (0) | Mild (1) | Moderate (2) | Severe (3) |
| Appropriateness |  |  |  |  |
| Intensity |  |  |  |  |
| Variability/Mobility |  |  |  |  |
| Range |  |  |  |  |
| Reactivity |  |  |  |  |
| **Affect Total (0-15 points)** |  | | | |
| Angry |  |  |  |  |
| Euphoric |  |  |  |  |
| Apathetic |  |  |  |  |
| Dysphoric |  |  |  |  |
| Apprehensive |  |  |  |  |
| **Mood (0-15 points)** |  | | | |
| **Domain II (0-30 points)**  **Mood + Affect** |  | | | |

| **Domain III: Cranial Nerves** | | | | | |
| --- | --- | --- | --- | --- | --- |
|  | Normal (0) | Mild (1) | Moderate (2) | Severe (3) | Bilateral (x2) |
| II: Optic |  |  |  |  |  |
| III: Oculomotor |  |  |  |  |  |
| IV: Trochlear |  |  |  |  |  |
| V: Trigeminal |  |  |  |  |  |
| VI: Abducens |  |  |  |  |  |
| VII: Facial |  |  |  |  |  |
| VIII: Auditory/Vestibular |  |  |  |  |  |
| IX: Glossopharyngeal |  |  |  |  |  |
| X: Vagus |  |  |  |  |  |
| XI: Accessory |  |  |  |  |  |
| XII: Hypoglossal |  |  |  |  |  |
| Nystagmus |  |  |  |  |  |
| Total (0-69): |  | | | |  |

| **Domain IV: motor function** | | | | | |  |
| --- | --- | --- | --- | --- | --- | --- |
|  | Muscle Bulk | Muscle strength | Muscle tone | Type: spasticity/rigidity/mixed | Muscle strength | Reflexes:  0 - normal  1 – increased without clonuses or decreased  2 - increased with clonuses or absent |
|  | 0 – Normal, 1-Mild; 2-Moderate, 3 Severe | | | | |  |
| Right upper Extremity |  |  |  |  |  |  |
| Left upper Extremity |  |  |  |  |  |  |
| Right upper Extremity |  |  |  |  |  |  |
| Left upper Extremity |  |  |  |  |  |  |
|  | | Normal (0) | | Mild (1) | Moderate (2) | Severe (3) |
| Plantar response (right) | |  | |  |  |  |
| Plantar response (Left) | |  | |  |  |  |
| **Cerebellar functions:** | | | | | | |
| Finger to nose  (right) | |  | |  |  |  |
| Finger to nose  (left) | |  | |  |  |  |
| Rapid alternating movements  (right) | |  | |  |  |  |
| Rapid alternating movements  (left) | |  | |  |  |  |
| Heel to shin  (right) | |  | |  |  |  |
| Heel to shin  (left) | |  | |  |  |  |
| **Cerebellar function total (0-18 points):** | |  | | | | |
| Resting tremor | |  | |  |  |  |
| Action Tremor | |  | |  |  |  |
| Postural tremor | |  | |  |  |  |
| Hypomimia | |  | |  |  |  |
| Gait | |  | |  |  |  |
| Gait description: | |  | | | | |
| **Motor total (0-85)** | |  | | | | |

| **Domain V: sensory** | | | |
| --- | --- | --- | --- |
| Location: | Normal – 0  Abnormal - 1 | Modality | Normal – 0  Abnormal - 1 |
| Stocking |  | Light touch |  |
| Stocking/glove |  | Pain and temperature |  |
| Dermatome |  | Vibration |  |
| Sensory nerve |  | Proprioception |  |
| Other |  | Other |  |
| Graphesthesia | | | |
| Normal (0) | Mild (1) | Moderate (2) | Severe (3) |
| Stereognosis | | | |
| Normal (0) | Mild (1) | Moderate (2) | Severe (3) |
| **Sensory total (0-16 points)** |  | | |

| **Other (0-5 points)** | | | |
| --- | --- | --- | --- |
| Frontal lobe release/other reflexes. Absent – 0, Present -1 | | | |
| Palmomental | Grasp reflex | Glabellar | Abnormal rate of blinking |
| Primitive reflexes. Absent – 0, Present -1 | | |  |

| Cognition (0-39 points) |
| --- |
| Affect and mood (0-30 points) |
| Cranial nerves (0-69 points) |
| Motor (0-85 points) |
| Sensory (0-16 points) |
| Other (0-5 points) |
| **Total 0-244 points** |
